# Research on Recognition Method of COVID-19 Images Based on Deep Learning

**DOI:** 10.1101/2020.12.09.20246371

**Authors:** Dongsheng Ji, Yanzhong Zhao, Zhujun Zhang, Qianchuan Zhao

**Affiliations:** School of Computer and Communication, Lanzhou University of Technology, LanZhou,China; Intelligence and Network Information Center, Tsinghua University, Beijing, China

**Keywords:** small sample, image recognition, deep learning, model fusion

## Abstract

In view of the large demand for new coronary pneumonia covid19 image recognition samples,the recognition accuracy is not ideal.In this paper,a new coronary pneumonia positive image recognition method proposed based on small sample recognition. First, the CT image pictures are preprocessed, and the pictures are converted into the picture formats which are required for transfer learning. Secondly, perform small-sample image enhancement and expansion on the converted picture, such as miscut transformation, random rotation and translation, etc.. Then, multiple migration models are used to extract features and then perform feature fusion. Finally,the model is adjusted by fine-tuning.Then train the model to obtain experimental results. The experimental results show that our method has excellent recognition performance in the recognition of new coronary pneumonia images,even with only a small number of CT image samples.

## 1. Introduction

As of July 30, 2020, there have been 17,039,160 people with new coronary pneumonia in the world, and it is growing at a rate of close to 300,000 per day. ^[1]^ In the diagnosis and treatment plan of the “New Coronavirus Infection Pneumonia Diagnosis and Treatment Plan (Trial Seventh Edition)” issued by the National Health and Medical Commission, in addition to the incubation period 1-14 days isolation observation based on epidemiological investigations, pneumonia symptoms based on clinical manifestations and the characteristics of the signs of illness, the results of laboratory nasopharyngeal swabs and nucleic acid testing of Yin/Yang, and effective oxygen therapy combined with antiviral and antimicrobial therapy, the most important diagnostic criterion is chest imaging. Based on the accumulation of clinical experience in the fight against new coronary pneumonia and the in-depth related research, some false negatives in the new coronavirus nucleic acid test may be questioned, especially for those with a negative nucleic acid test and typical CT findings, which may affect clinical investigation and treatment. ^[2]^ On February 4, 2020, the National Health Commission released the latest “New Coronavirus Pneumonia Diagnosis and Treatment Plan” (Fifth Edition) to update the fourth version. While confirming the diagnosis and paying attention to the imaging findings, the patients CT imaging findings are also included in the discharge criteria, and imaging findings are used as the basis for clinical suspicion and diagnosis in Hubei. ^[3]^ Compared with images of other modalities, CT images have higher definition and resolution. Therefore, chest imaging examination is an important medical imaging method for timely identification of COVID-19 cases. Through the diagnosis of chest imaging, medical staff can grasp the imaging characteristics of COVID-19 cases more accurately, such as early appearance of multiple small patchy shadows and interstitial changes, which are obvious outside the lung. Then,it develops into multiple ground-glass shadows and infiltration shadows in both lungs. In severe cases, lung consolidation and pleural effusion are rare. It has important guiding value for accurately estimating the condition and its development, formulating treatment plans and evaluating prognosis.

This article uses the domain knowledge obtained from the detection and recognition of medical imaging targets to assist doctors in the diagnosis and treatment of COVID-19 disease, which is conducive to the early detection of lung lesions, etc., especially telemedicine and diagnosis, which has very important application value and Academic significance. The intervention of the application of computer intelligent image analysis technology provides new ideas and ways to accurately identify and assist medical diagnosis of COVID-19 disease. At the same time, it also has a high reference value for the further study of intelligent imaging-assisted diagnosis technology for human severe viral pneumonia diseases and other newly emerging respiratory syndrome diseases.

### 2 Relevant work

Covid19’s image recognition can be regarded as image recognition. Image recognition mainly relies on pattern recognition, and current machine learning is developed from pattern recognition. Changes in common chest CT ground-glass features of new coronary pneumonia are the most common manifestations of COVID-19 infection. The CT manifestations of pneumonia caused by new coronavirus infection are diverse, with ground-glass shadows and actuality becoming dominant, lacking specificity. Generally, the clinical symptoms are mild and the lung lesions are few, which often brings certain difficulties to the diagnosis. For suspected cases, CT examination finds abnormal manifestations in the lungs, which can assist in early treatment and intervention. However, the diagnosis requires a comprehensive judgment based on epidemiological history, new coronavirus testing and radiological manifestations.

On February 28th, a research team led by Academician Nanshan Zhong (37 experts from the China New Coronary Pneumonia Medical Expert Group, research units include the National Respiratory Disease Clinical Research Center, Wuhan Jinyintan Hospital and other 9 authoritative institutions) in the New England Journal of Medicine (NEJM) published the research results on the “Clinical Features of China’s 2019 Novel Coronavirus Disease”, ^[4]^ mainly related to patient symptoms, confirmed patients, clinical symptoms and infection signs of suspected patients. From the perspective of imaging evaluation, chest X-ray or CT image was analyzed for lesion characteristics. The research team used detailed data to compare and analyze the characteristics of SARS, MERS and seasonal influenza cases, showing that the new coronary pneumonia has different trends. Domestic scholars wrote an article in the “Nature” magazine on the study of the characteristics of new coronary pneumonia, and discussed the clinical characteristics and imaging detection characteristics. ^[5]^ Researchers from the National Supercomputing Center of Sun Yat-sen University used the parallel computing deep learning method to study the characteristics of COVID-19 imaging lesions^.[6]^ The training model can locate the main regional features, especially ground glass shadows, and the model can be fast. The research team of Central South University studied the short-term disease progression and clinical characteristics of patients with new coronary pneumonia, ^[7]^ carried out characteristic studies on chest CT images of confirmed cases, analyzed the severity of the disease, and made clinical treatment recommendations.

Doctors from the Radiology Department of Shanghai Public Health Clinical Center, together with researchers from the Shanghai United Imaging Research and Development Department and doctoral students of Shanghai University, used deep learning to build an automatic segmentation and quantification system based on deep learning, ^[8]^ mainly used image segmentation theory for research The chest CT infection area and the overall structure of the lungs are annotated for each case using the method of man-machine loop optimization. Researchers from East China Normal University, the Key Laboratory of Artificial Intelligence of the Ministry of Education, Ryerson University in Canada, and Shanghai Jianglai Data Technology Co., Ltd. used depth cameras and deep learning to study human abnormal breathing patterns, which are clinically significant for 6 types Breathing patterns are classified to help large-scale screening of patients infected with COVID-19. ^[9]^ Researchers from Huazhong University of Science and Technology and Wuhan University of Technology used machine learning models to predict the criticality of patients with severe COVID-19, and identify three main clinical features,assessing the risk of death accurately and quickly,which has important clinical significance. ^[10]^

Dr. Shuai Wang from the Cancer Hospital of Tianjin Medical University used deep learning methods to extract COVID-19 image features, established a learning model to analyze positive cases, and provided a theoretical basis for timely and accurate diagnosis of COVID-19. ^[11]^ Experts from the Affiliated Hospital of Huazhong University of Science and Technology used three-dimensional CT to detect new coronary pneumonia, and a three-dimensional neural network based on weakly supervised deep learning to classify positive and negative cases to quickly identify COVID-19 cases. ^[12]^ Researchers such as Asmaa and Mohammed from Arthurs University and Birmingham City University, ^[13]^ In view of the high availability of COVID-19 annotated image data sets, use convolutional neural networks to identify and classify new crown images, and use the class decomposition mechanism to study its class boundary to deal with the irregularities in the data set, and having great performance.

On May 10th, the Fei Shan team developed a deep learning (DL)-based segmentation system to automatically quantify the infected region of interest (ROIS). The performance of the system was evaluated by comparing the automatic segmentation of the infected area on the chest CT of 300 patients with new coronary pneumonia and the manual delineation of the infected area. For the rapid manual drawing of training samples and manual intervention of automatic results, the artificial loop (HITL) strategy is used to assist radiologists in segmenting the infected area, which greatly reduces the total segmentation time after the model is updated three times to 4 minutes. The average estimation error (POI) of the whole lung infection rate (POI) was 0.3%. Finally, possible applications are discussed, including but not limited to the analysis of subsequent CT scans and the distribution of infections in lobes and segments associated with clinical manifestations. ^[14]^On May 24, the Ezz El-Din Hemdan team introduced a new deep learning framework, the COVIDX network, to help radiologists automatically diagnose new coronary pneumonia in X-ray images. COVIDX-Net includes seven different deep convolutional neural network models, such as the improved visual geometry group network (VGG 19) and the second version of Google MobileNet. Each deep neural network model can analyze the normalized intensity of X-ray images to classify the negative or positive cases of new coronary pneumonia. ^[15]^ On May 27th, Jianpeng Zhang’s team distinguished viral pneumonia from non-viral pneumonia and healthy controls into a category-based anomaly detection problem, and proposed a shared feature extractor, anomaly detection module, and a confidence prediction module It consists of a trust-aware anomaly detection (CAAD) model. If the abnormality score generated by the abnormality detection module is large enough, or the confidence score estimated by the confidence prediction module is small enough, the input is regarded as an abnormal case (ie, viral pneumonia). ^[16]^On May 31, the Muhammad Farooq team established an open source and open access data set, and proposed an accurate convolutional neural network framework to distinguish 19 COVID cases from other pneumonia cases. They proposed a multi-class classification model of three different infection types and normal people with high computational efficiency and high accuracy. This model helps to screen 19 COVID cases early and helps reduce the burden on the medical system. ^[17]^In addition, other scholars have also conducted in-depth research in the field of deep learning-assisted diagnosis of covid19. For example, Boyi Liu’s team proposed the use of federated learning methods to train and deploy experiments on new coronary pneumonia data to verify the effectiveness of the method. They also compared the performance of four popular models (MobileNet, Resnet 18, MbedeNet, and COVID-net) with and without federated learning frameworks. Their work aims to inspire more research on federal learning about COVID-19. ^[18]^ The Minaee team performed transfer learning on a subset of 2000 radiographs and trained four popular convolutional neural networks, including Resnet 18, Resnet 50, SqueezeNet and DenseNet-121, in order to analyze the chest X-ray images To identify new coronary pneumonia. They evaluated these models on the remaining 3,000 images, most of which have a sensitivity of 97% and a specificity rate of about 90%. ^[19]^The Arman Haghanifar team collected a large number of chest X-ray images from various sources and prepared the largest publicly accessible data set. Finally, using the migration learning paradigm, the famous CheXNet model was used to develop COVID-CXNet. This model is based on relevant and meaningful features, has accurate positioning capabilities, and can detect new coronavirus pneumonia. ^[20]^The Kumar team used new and up-to-date data to improve the recognition of a global deep learning model, and learned from these data to improve the recognition ability of new coronary pneumonia patients based on CT slices. In addition, the combination of blockchain and federated learning technology can collect data from different hospitals without leaking data privacy. First, they collected real life data of patients with new coronary pneumonia and opened it to the research community. Second, they used various deep learning models (VGG, DenseNet, AlexNet, MobileNet, ResNet, and Capsule Network) to screen patients with new coronary pneumonia. Identify these patterns. Third, the method of combining joint learning and block chain is adopted to realize the safe sharing of data between hospitals. In the end, their results show that patients with new coronary pneumonia are better tested. ^[21]^

In 2012, AlexNet was proposed by Alex Krizhevsky and others set off a wave of deep learning. The error rate of the network in the ImageNet Challenge was reduced by more than 10% compared with the previous championship, and 10.8 percentage points higher than the runner up. ^[22]^ In the same year, Feifei Li of Stanford University and others completed the imagenet data set, which provided the possibility of migration learning for image recognition. In 2014, Adam was widely used due to its easy to fine-tune features. It is based on the idea of adapting to different learning rates for each parameter. The function used in this model fusion is Adam. ^[23]^ VGG was proposed by the Visual Geometry Group of the Department of Science and Engineering, University of Oxford in 2014. The main work is to prove that increasing the depth of the network can affect the final performance of the network to a certain extent. Compared with AlexNet in 2012, a high advance of VGG is to use continuous 3×3 small convolution kernels to replace the larger ones in AlexNet (AlexNet uses 11×11, 7×7 and 5×5 convolution kernels). The main effect is to increase the depth of the network. ^[24]^ In 2015, with the emergence of ResNet, the performance of neural networks on visual classification tasks surpassed humans for the first time. ^[25]^ Christian Szegedy then made the Inceptionv2 model, which is implemented as inceptionV3 in the code. This model can balance the depth and width of the network with limited computing resources. ^[26]^ In 2016, DenseNet was proposed.The authors start from the feature, and achieve better results and fewer parameters through the extreme use of the feature. The central idea is to directly connect all layers under the premise of ensuring the maximum information transmission between layers in the network. One advantage of DenseNet is that the network is narrower and has fewer parameters. ^[27]^ In 2017, Xception improved the classification performance on ImageNet based on InceptionV3. ^[28]^ In 2018, a more efficient NasNet model appeared. ^[29]^

This paper applies deep convolutional neural network to realize image feature detection of COVID-19 cases. Through the investigation of the literature and the study of the theory of deep convolutional neural networks, a small-sample deep learning-based image recognition model for new coronary pneumonia positive images was designed. First, the sample CT images are processed using the latest small sample image preprocessing method, which greatly expands the training samples. Then, the model that screened the experimental model that helped improve the recognition accuracy was feature extraction and fusion, which greatly improving the accuracy of training. Through the fine-tuning of the fusion model, the accuracy of model training can be further improved. Finally, train the model, use matplotlib and seaborn tools to display the model and experimental results, and evaluate the training accuracy, verification accuracy, training loss, and verification loss of the model. Experiments show that the recognition accuracy of the model proposed in this article is as high as 96%, especially in the condition of a small number of samples of new coronary pneumonia images.

## 3 Based on small sample deep learning detection model

This paper uses a small sample-based image enhancement method and fine-tunes and merges multiple models of deep learning to further improve the recognition rate of positive image detection.

### 3.1 Image recognition convolutional neural network and loss function optimization

Convolutional neural network is one of the representative algorithms of deep learning, and image recognition based on deep learning also uses convolutional neural network. The convolutional neural network is characterized by its ability to characterize learning and translation invariance. Compared with traditional image recognition algorithms, it has a considerable breakthrough, and its recognition accuracy is greatly improved.

The loss function is a function used to calculate the difference between the label value and the predicted value. It must be used when the deep learning model is compiled. The loss function is a cost function defined on the training set.

Cross entropy cost function:

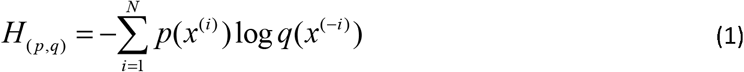

The two-category cost function used this time is:

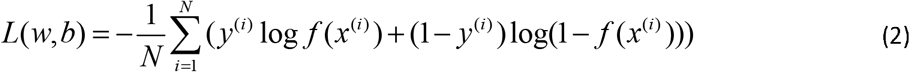

Where f(x) can be a sigmoid function. Or other activation functions in deep learning. And y^(i)^□0,1

The optimizer is also one of the parameters that must be used during deep learning compilation. It is used to update and calculate the network parameters that affect model training and model output, so that it approximates or reaches the optimal value, thereby minimizing (or maximizing) the loss function. The optimizers provided by Keras are BGD, SGD, MBGD, Momentum, NAG, Adagrad, Adadelta, RMSprop, Adam. This experiment mainly uses rmsprop and adam optimizers. The relationship between the loss function and the optimizer balances the detection quality and minimizes the loss of image features.

### 3.2 Image enhancement of small samples

In the past, deep learning must use a large number of training samples as support, otherwise it is prone to overfitting during training. However, this experiment uses the patient’s ct image data. Due to the patient’s privacy problem, the amount of data is seriously insufficient, so a small sample of image enhancement technology is used to perform a large amount of data expansion. This experiment uses the latest keras image augmentation technology. Compared with the traditional image augmentation technology using pillow or opencv, this technology is simpler and faster. The optional expansion methods basically meet the needs,the generated pictures and the randomness is stronger.

The picture before extension is shown in Figure 1,and the picture after enlargement is shown in Figure 2.

**Figure 1.**
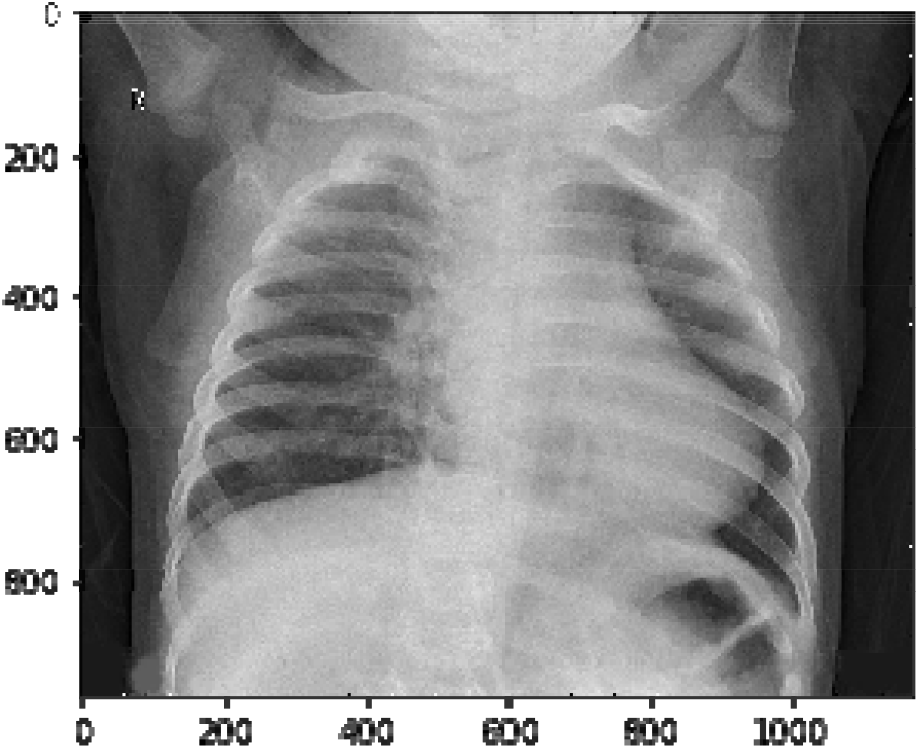
Before extension

**Figure 2.**
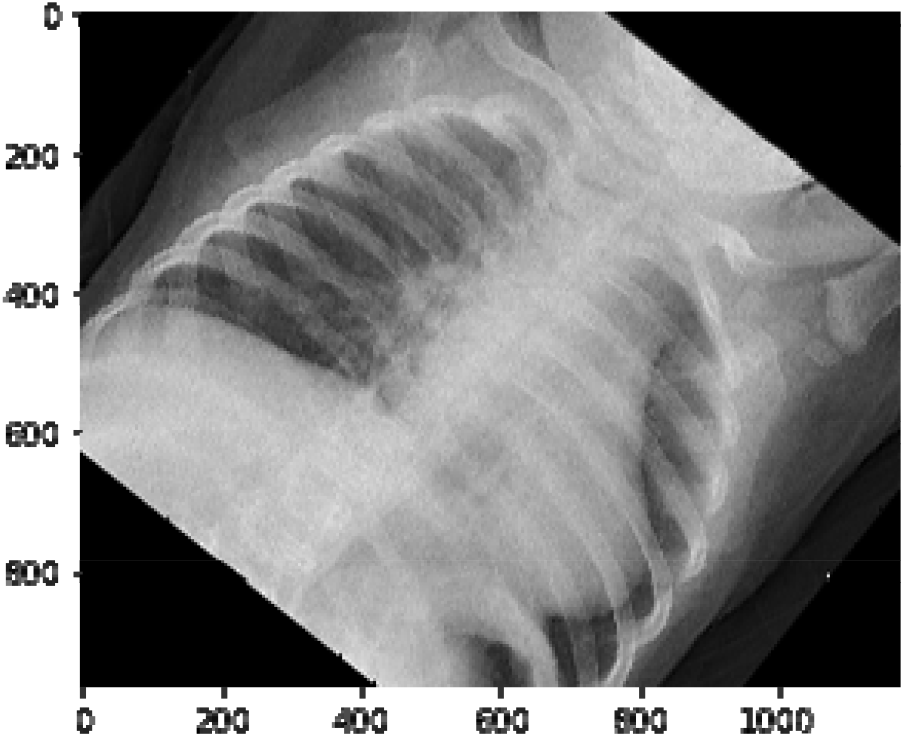
After enlargement

The augmentation technologies used this time include:

- Miscut transformation, the effect is to keep the x coordinate (or y coordinate) of all points unchanged, while the corresponding y coordinate (or x coordinate) is translated proportionally, and the magnitude of the translation and the point to the x axis (or y) Axi) is proportional to the vertical distance.
- Random rotation angle, by random rotation between (0, a certain maximum angle), a large number of random simulation pictures are generated.
- Horizontal position translation and up and down position translation, the translation distance is 0 to 0.5 times the length or width of the picture in this experiment.
- The picture is enlarged in the length and width directions, so that it can be zoomed in and out to the same degree in both the length and width directions.
- Randomly flip the image horizontally or vertically, and randomly select the image to flip each time.
- Fill in the missing parts of the picture. Keras has the effects of “reflect”, “wrap”, “nearest, and “constant”.

### 3.3 Model fusion

Model fusion is a method of combining a group of base classifiers in a certain way to improve the overall performance of the model. This model fusion is similar to the voting mechanism, that is, multiple model prediction results are voted, and the minority obeys the majority. Finally the hit probability of the majority is output.

The fusion model of Inception_v3, DenseNet, ResNet50, Xception, VGG19, NASNetLarge, and InceptionResNetV2 is used this time,as shown in Figure 3. These models have their own advantages: the Inception model increases the adaptability of the network to multiple scales.DenseNet improves the flow of information and gradients in the network, and reduces the number of parameters. Resnet proposes a residual structure and a bottleneck layer structure to solve the depth For the degradation problem of the network.Xception proposes a deep separable convolution operation, which reduces a large number of parameters. The VGG19 structure is relatively simple. A simple network can still obtain high accuracy. NASNetLarge uses a neural structure search framework to build Very high accuracy. InceptionResNetV2 is improved from InceptionV3, and its frame accuracy is higher than InceptionV3.

**Figure 3.**
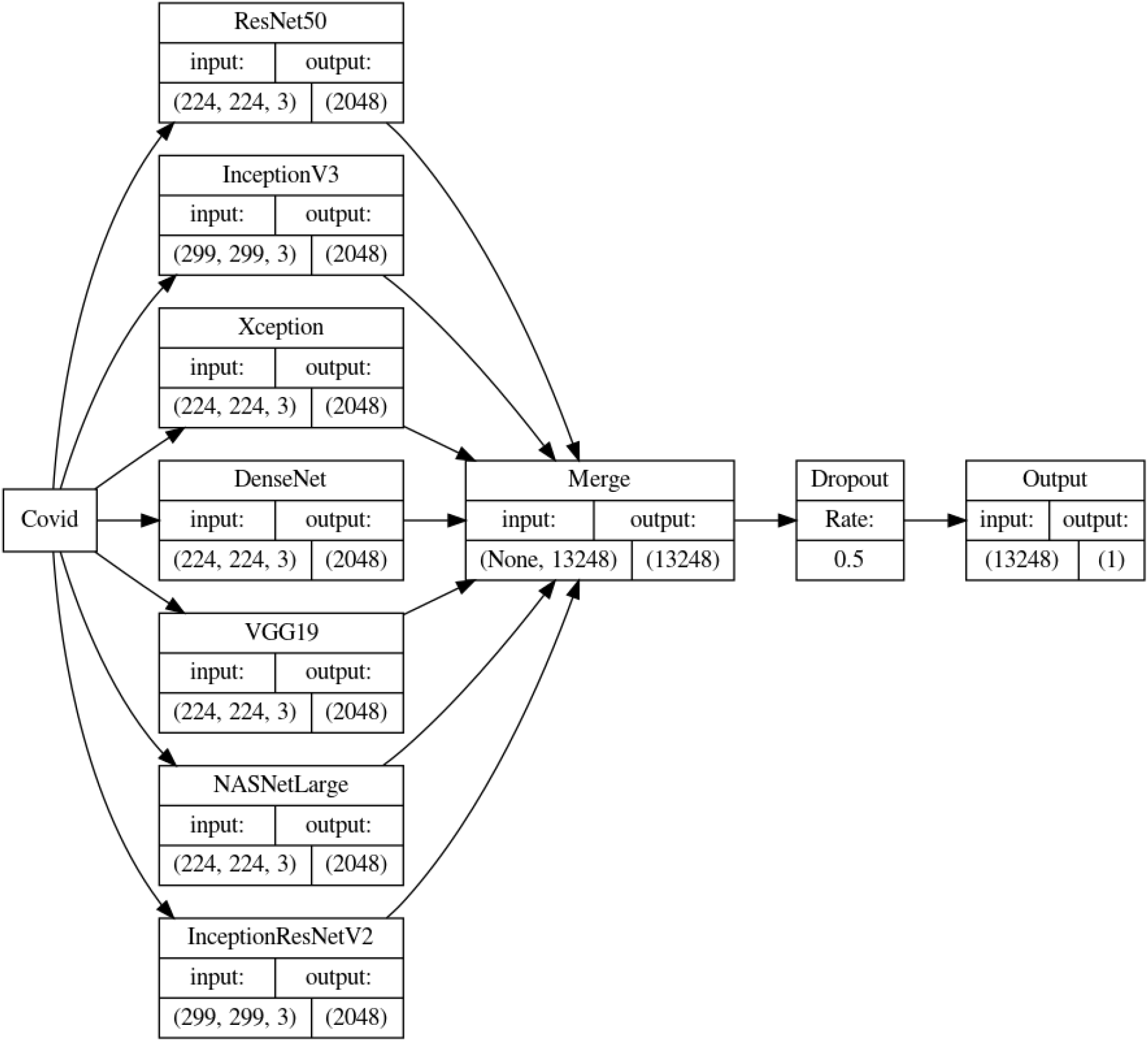
Model fusion diagram

## 4. Experimental process

### 4.1 Introduction to Data Set

The data set of this experiment comes from kaggle and github. The number of samples is 6000, and the training set is 2450. Data division is shown in Figure 4.

**Figure 4.**
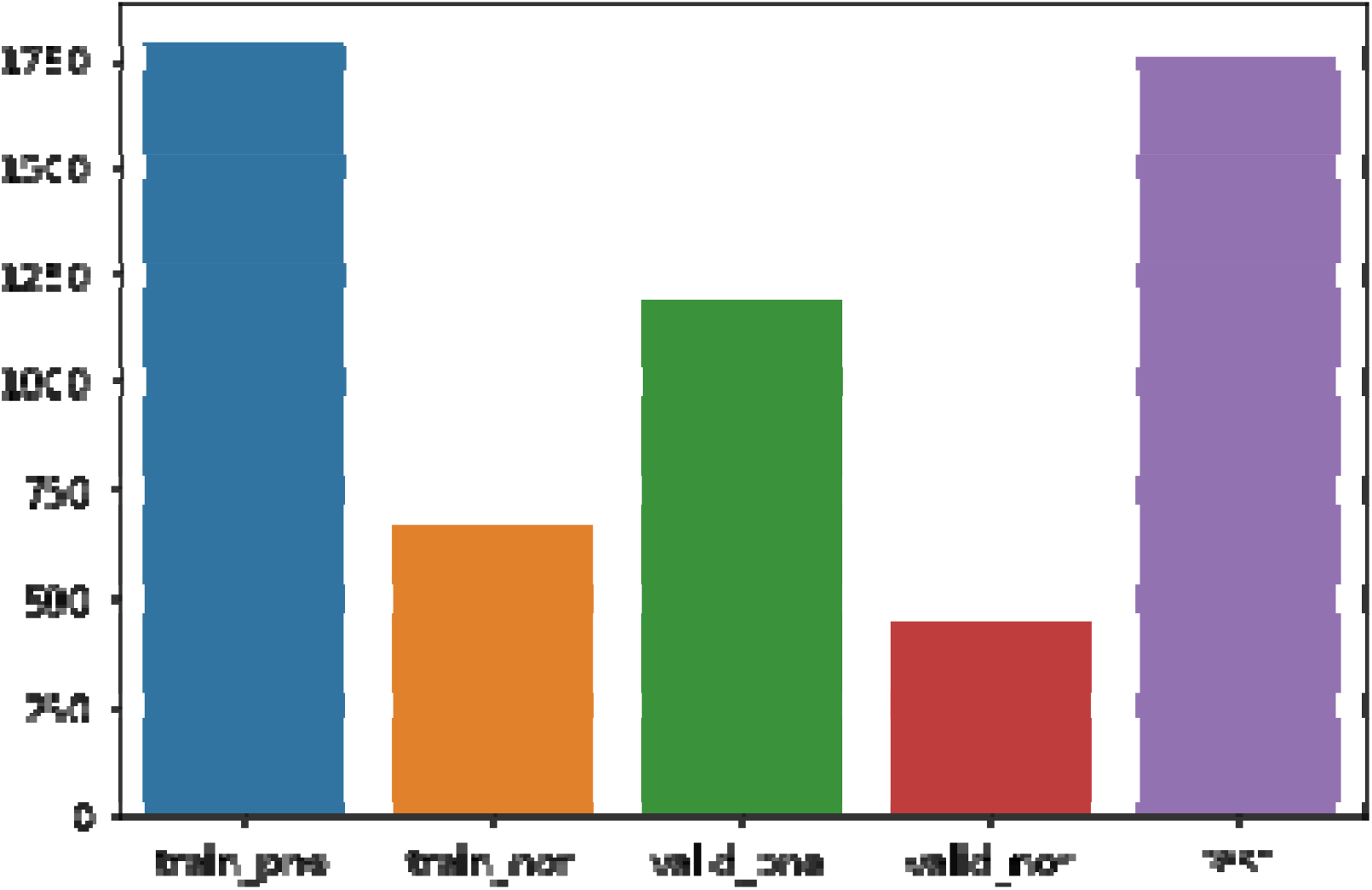
data set distribution

Legend: train_pne is the pneumonia training set, train_nor is the non-pneumonia training set, vaild_pne is the pneumonia validation set, vaild_nor is the non-pneumonia validation set, and test is the test set

### 4.2 Data preprocessing

Data preprocessing adopts image augmentation technology based on small samples, and scales each picture. The scale factor rescale of this style is 1./255

The size of the picture is adjusted according to the migration model, some are 224, some are 299, and the channels are all 3.

The Figure 5 is a processed normal image, and the Figure 6 is an image of a person infected with pneumonia.

**Figure 5.**
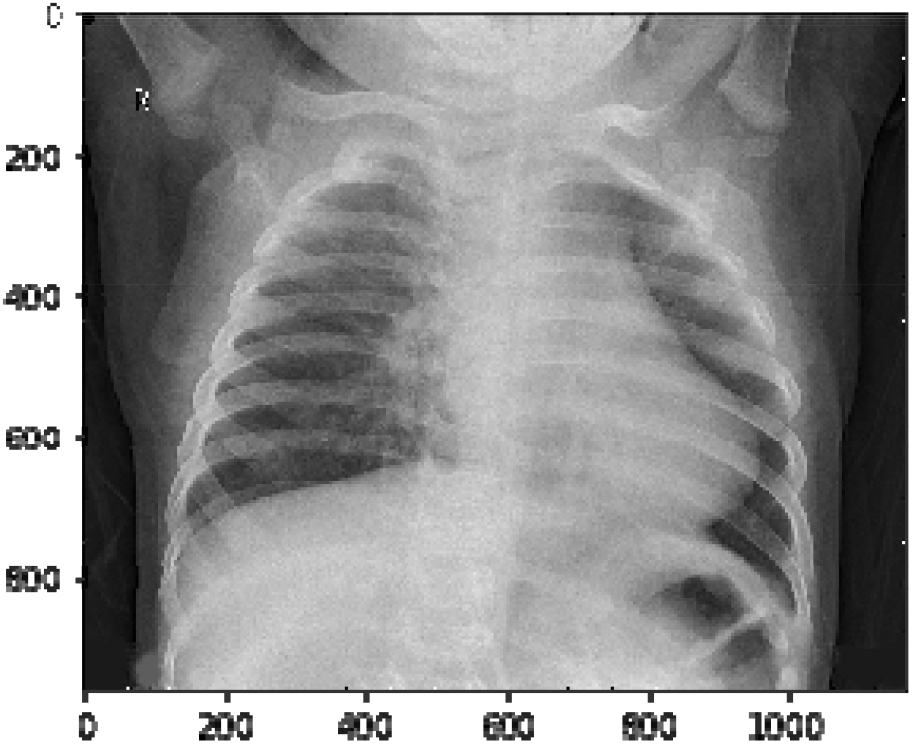
health image

**Figure 6.**
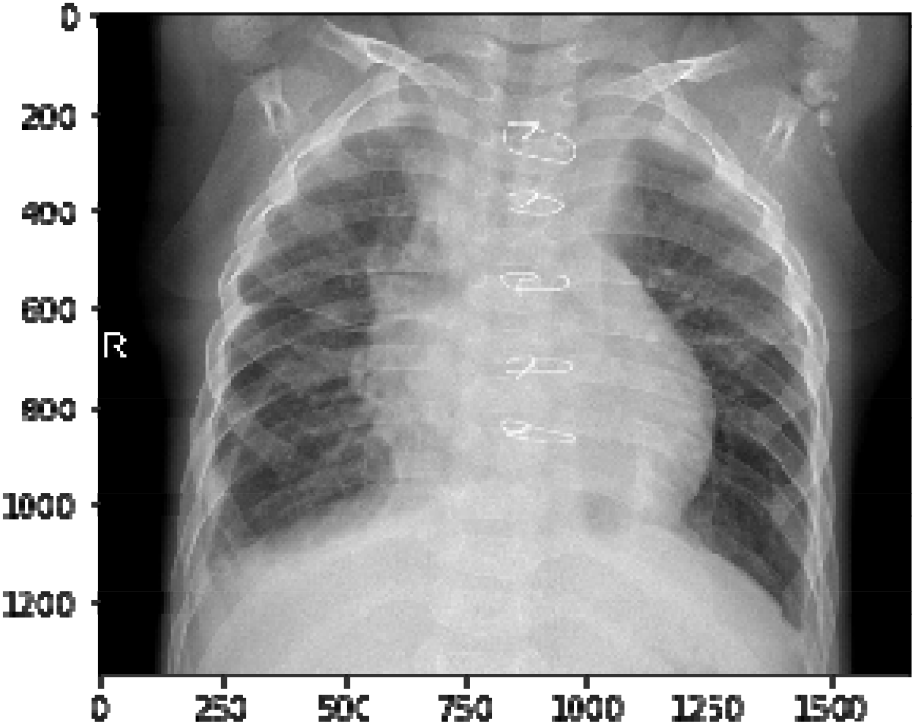
pneumonia image

### 4.3 Model Fusion

The fusion model of Inception_v3, DenseNet, ResNet50, Xception, VGG19, NASNetLarge, and InceptionResNetV2 is used this time,as shown in Figure 7. First download these official models and use these models as base models, and then use the gap layer to directly realize the regularization of the entire network structure and prevent overfitting. Second, two generators are used to generate a large amount of enhanced image data, and then the enhanced image data is used to derive the feature vector of each model. After that, the feature vector is fused through the np.concatenate function, and the model fusion is completed.

**Figure 7.**
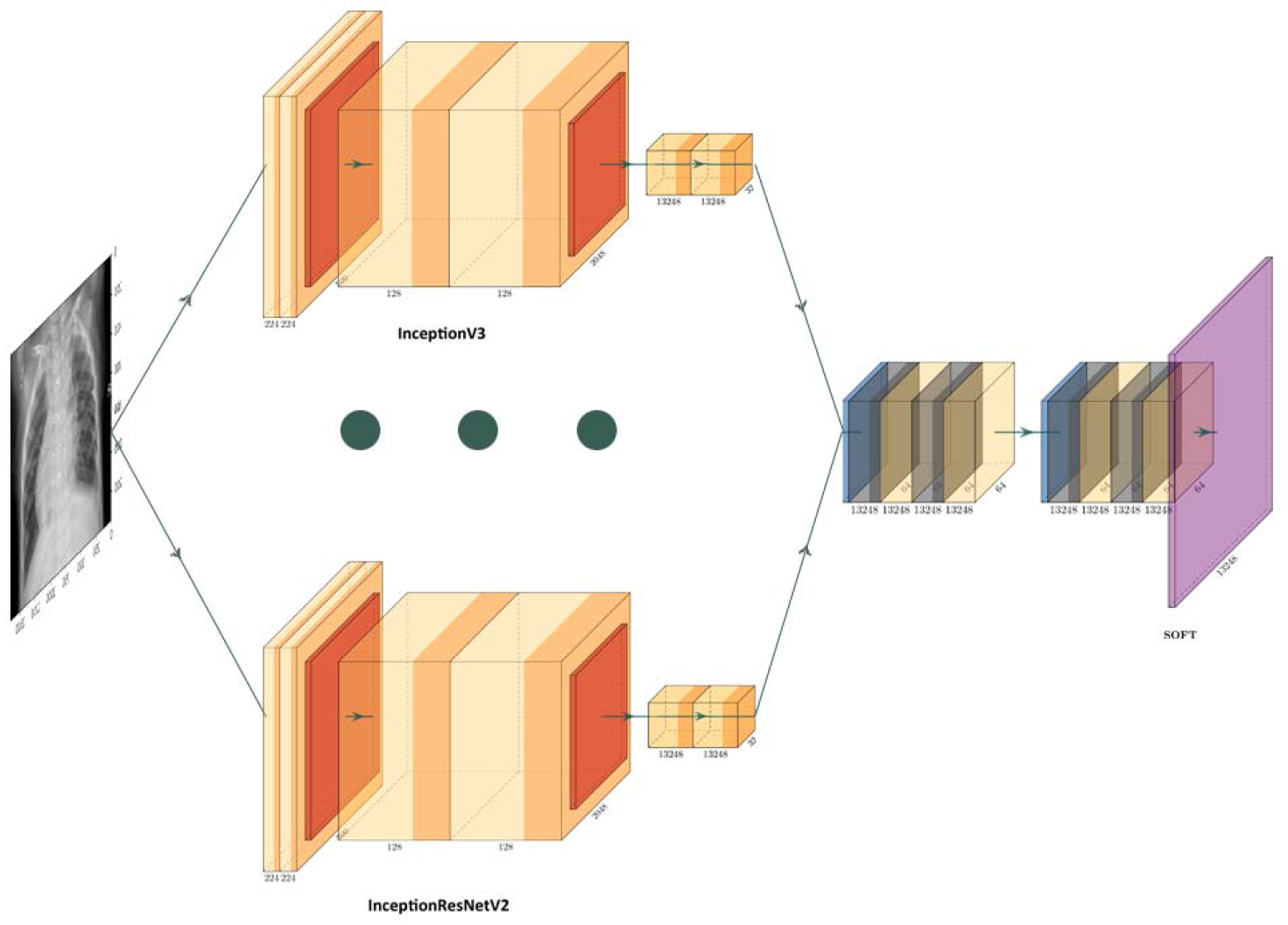
model fusion diagram

### 4.4 Model fine-tuning

Build a fine-tuning model class to properly fine-tune the final model to improve accuracy. After fine-tuning, dropout is used to randomly select some layers to prevent overfitting, and then perform model training

### 4.5 Machine configuration environment and model training

This training environment is kaggle, which is essentially a jupyternotebook page built by docker. The GPU used by kaggle is Nvidia Tesla P100-PCIE-16GB 1.3285GHz, the CPU is Intel(R) Xeon(R) CPU 2.3Ghz, and the memory is 14GB. The disk size is 5.2GB. The Aconda version is 3.6.6. Machine learning and deep learning modules such as tensorflow, keras, and sklearn are pre-installed. These modules are the latest version.

The final model training optimizer uses Adam, the learning rate is 0.00001, the loss function is binary_crossentropy, batch_size is 2, epochs is 100, and validation_split is 0.4

### 4.6 experimental results

Use the optimized DenseNet model test, the Figure 8 shows accuracy and the Figure9 shows loss.

**Figure 8.**
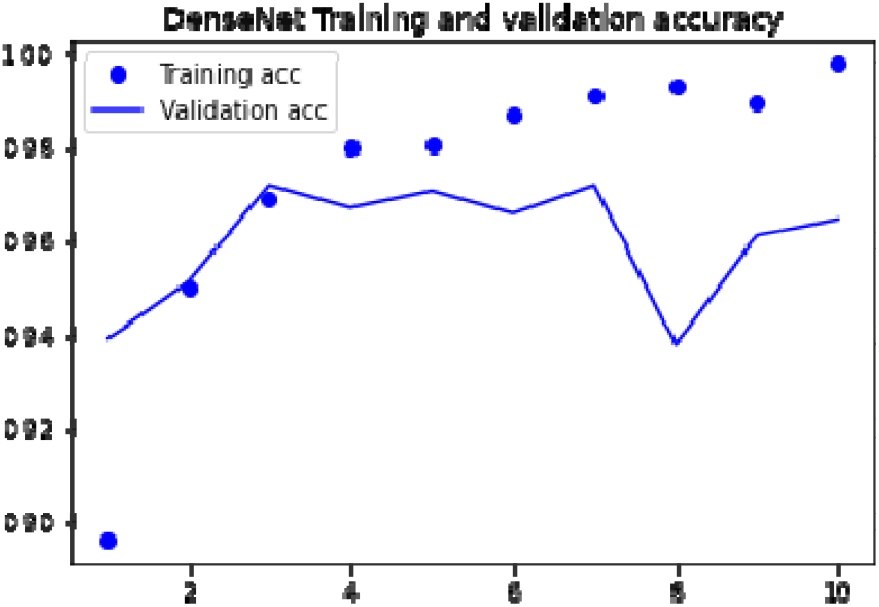
DenseNet accuracy

Use the optimized InceptionV3 model test,the Figure 10 shows accuracy and the Figure 11 shows loss.

**Figure 9.**
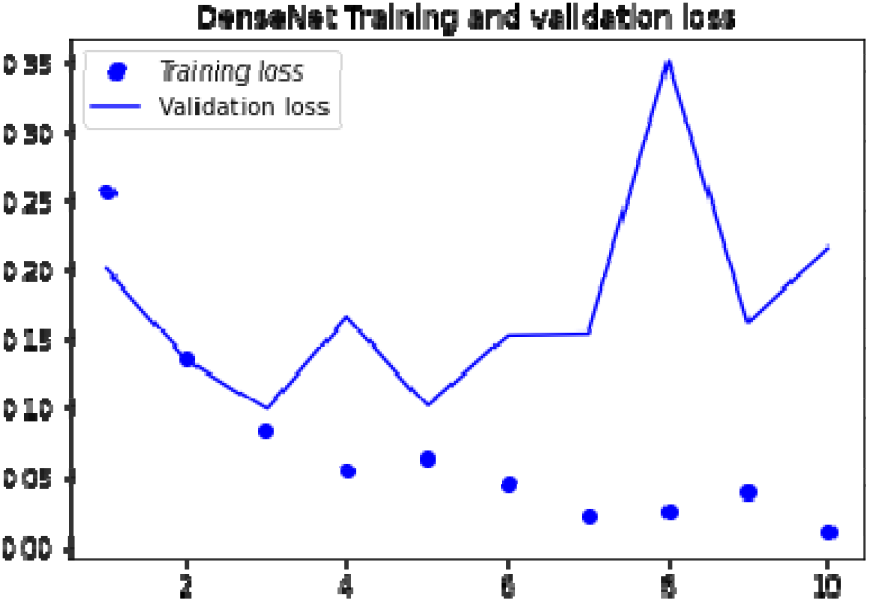
DenseNet loss

**Figure 10.**
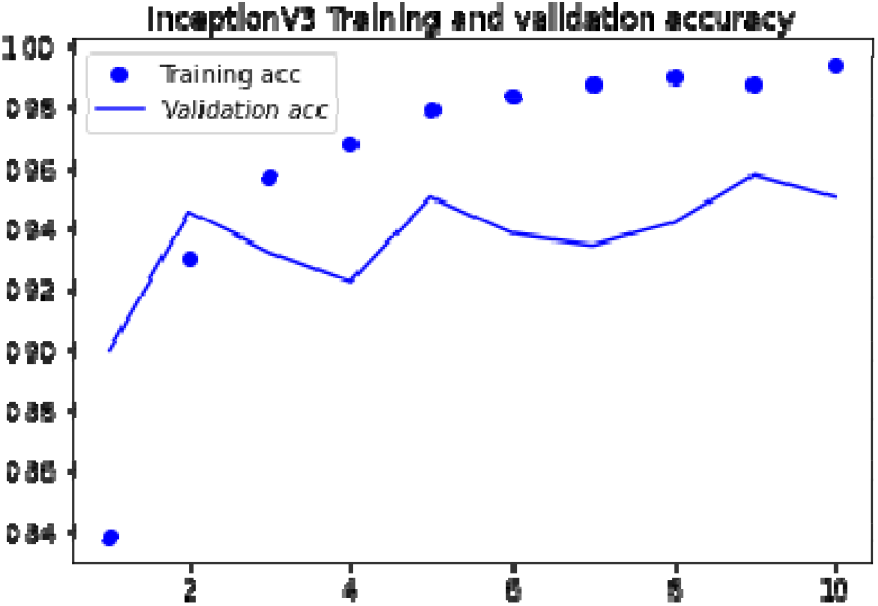
InceptionV3 accuracy

**Figure 11.**
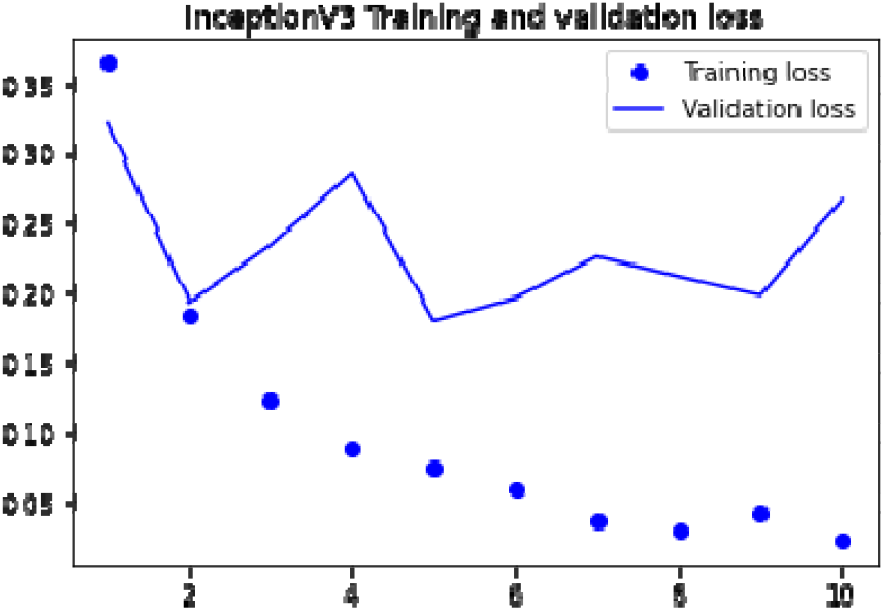
InceptionV3 loss

Use the optimized DenseNet model test, the Figure 12 shows accuracy and the Figure 13 shows loss.

**Figure 12.**
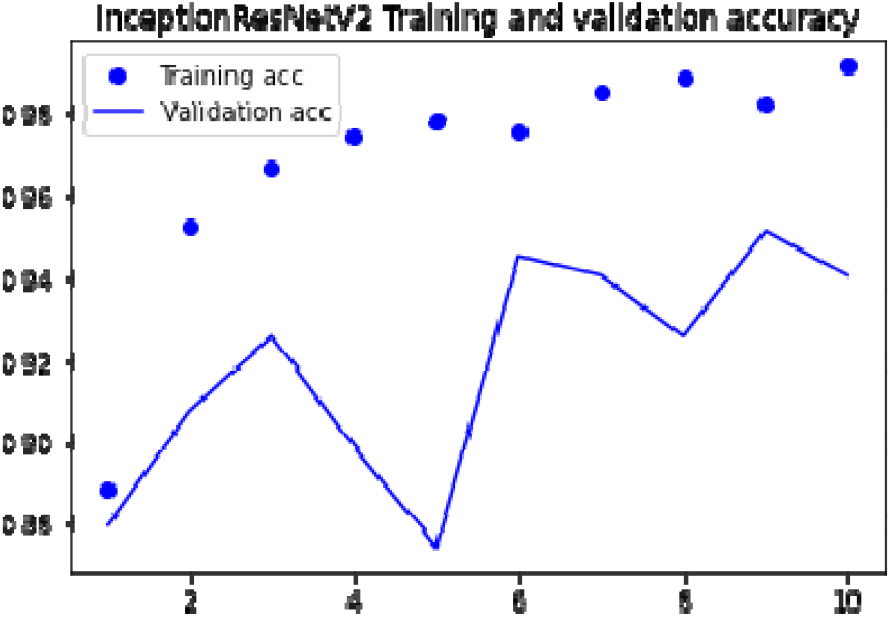
InceptionResNetV2 accuracy

**Figure 13.**
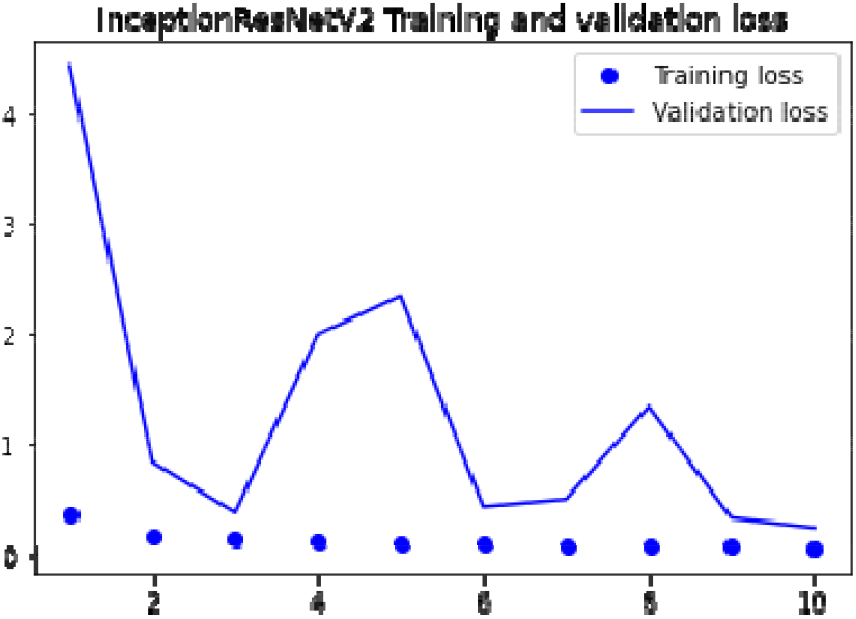
InceptionResNetV2 loss

Use the optimized DenseNet model test, the Figure 14 shows accuracy and the Figure 15 shows loss.

**Figure 14.**
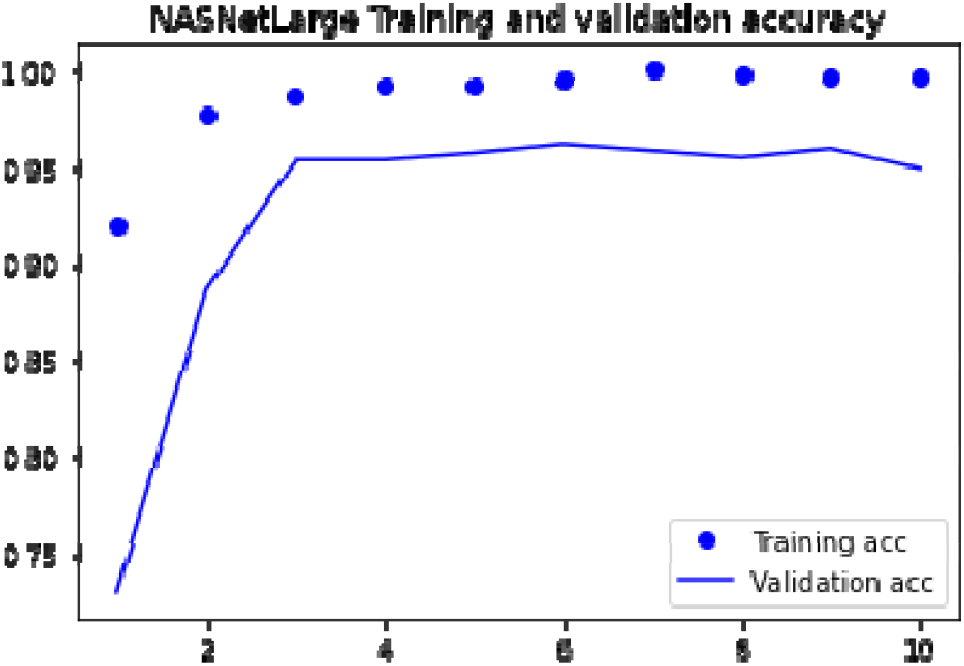
NasLargeNet accuracy

**Figure 15.**
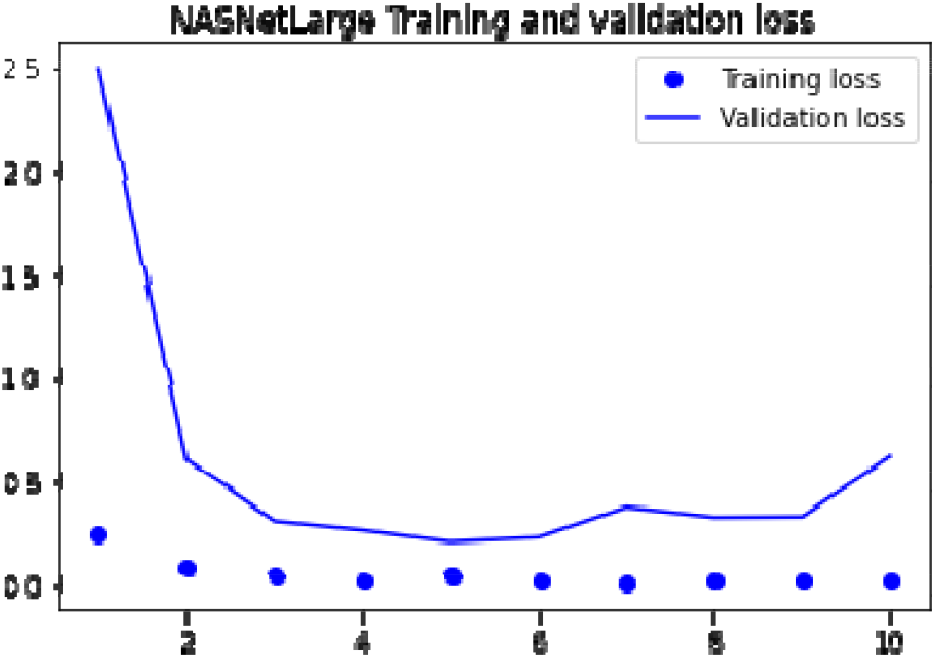
NasLargeNet loss

Use the optimized ResNet50 model test, the Figure 16 shows accuracy and the Figure 17 shows loss.

**Figure 16.**
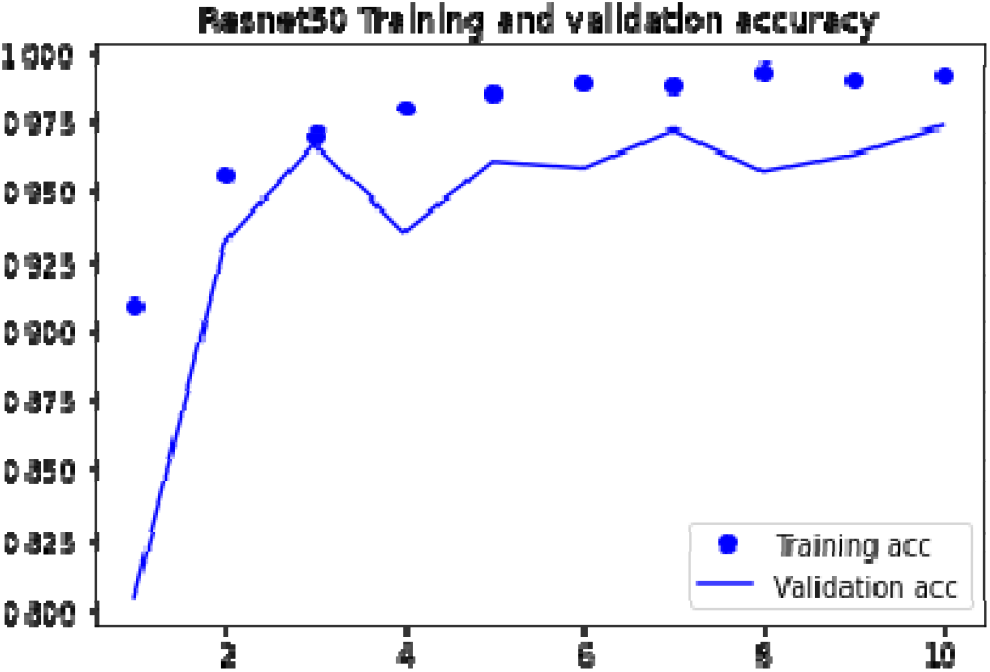
ResNet50 accuracy

**Figure 17.**
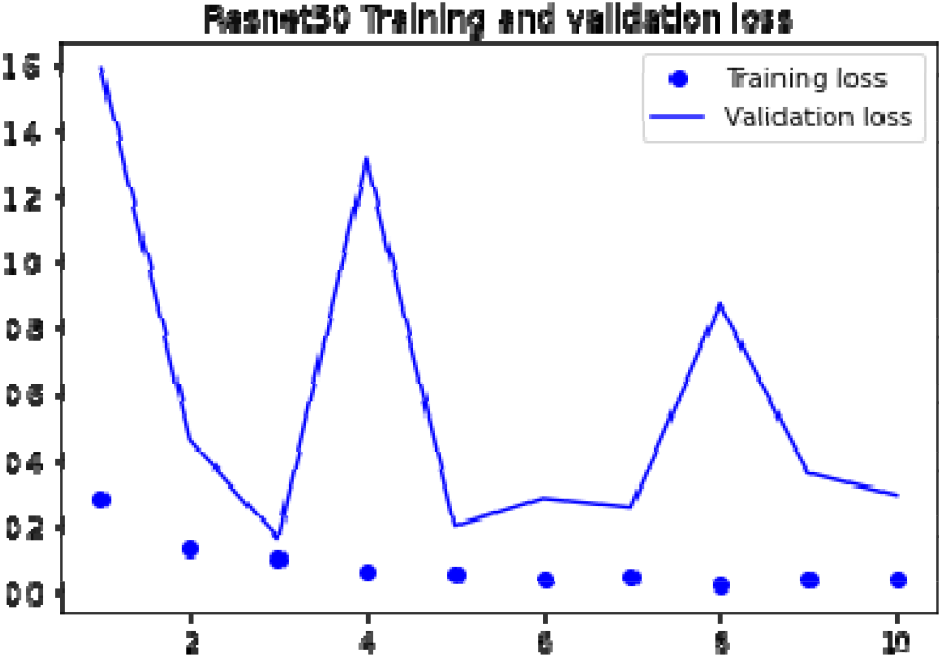
ResNet50 loss

Use the optimized VGG19 model test, the Figure 18 shows accuracy and the Figure 19 shows loss.

**Figure 18.**
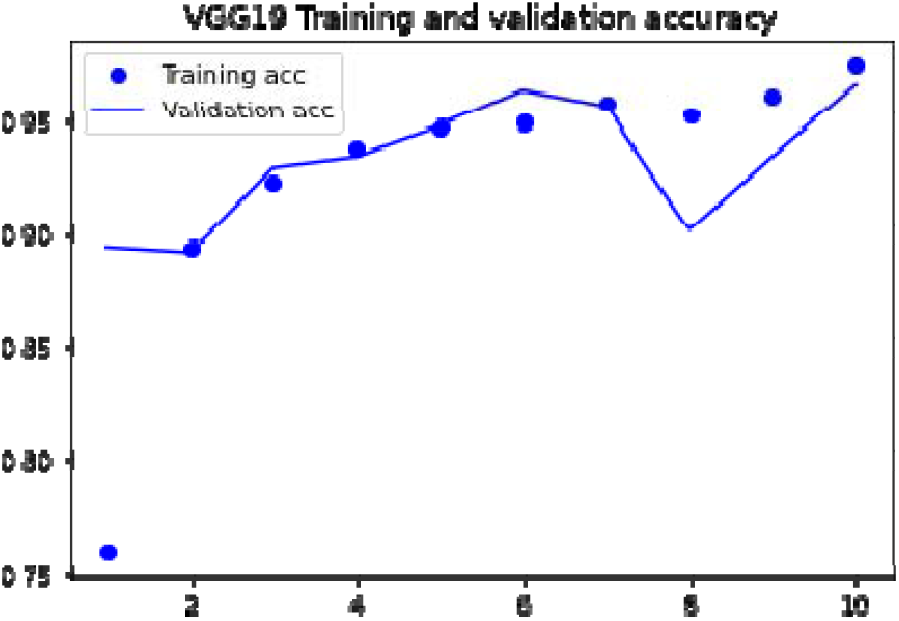
VGG19 accuracy

**Figure 19.**
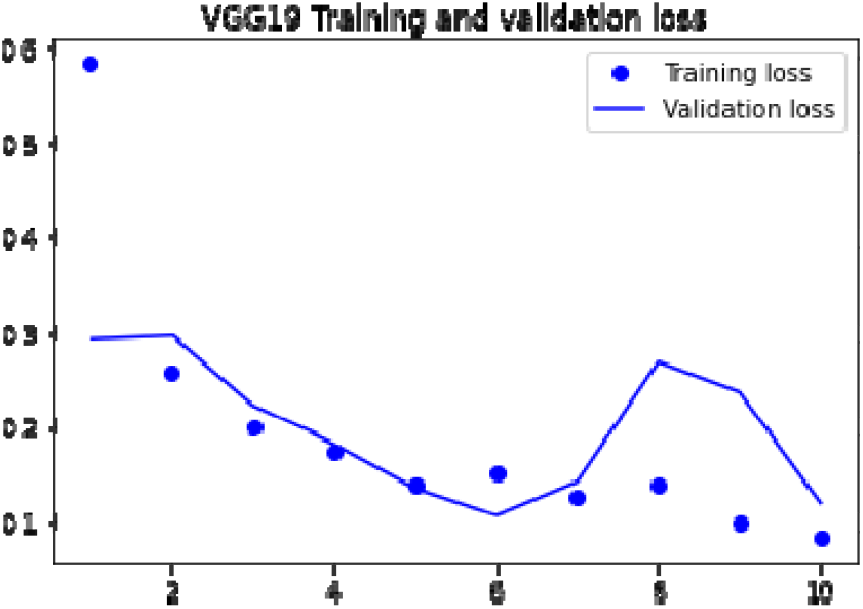
VGG19 loss

Use the optimized Xception model test, the Figure 20 shows accuracy and the Figure 21 shows loss.

**Figure 20.**
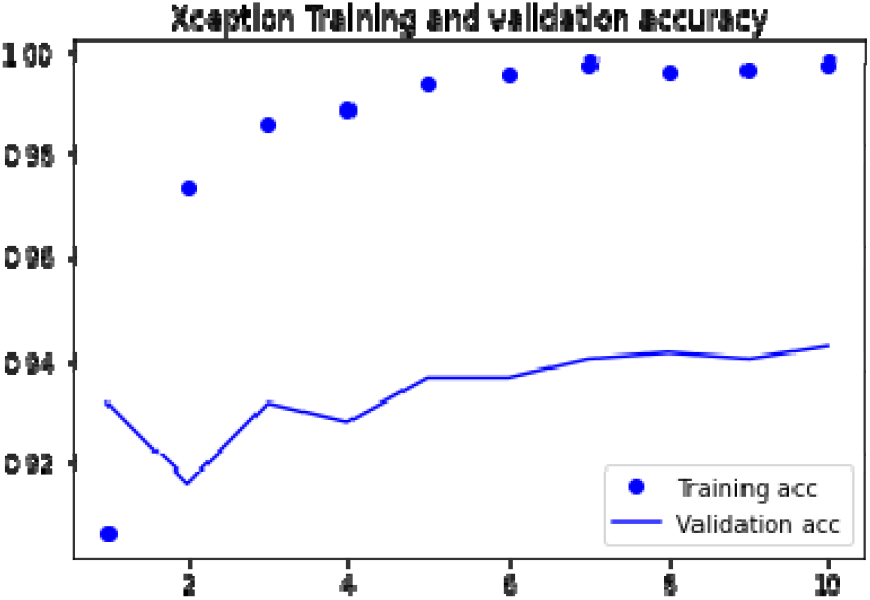
Xception accuracy

**Figure 21.**
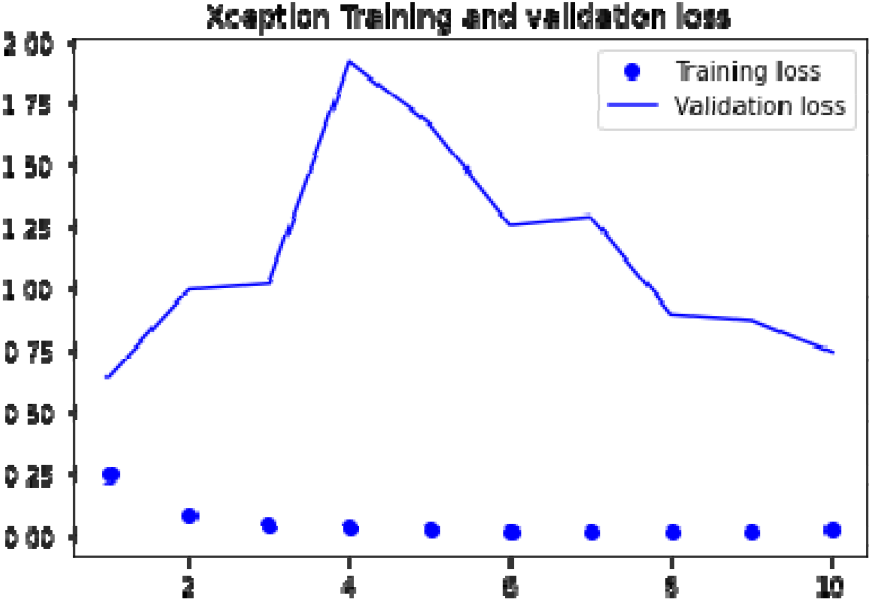
Xception loss

According to the final experimental results, the Figure 22 shows accuracy and the Figure23 shows loss,this training has neither over-fitting nor under-fitting. The accuracy of the final model is 96%, which has high reference value in the diagnosis of new coronary pneumonia.

**Figure 22.**
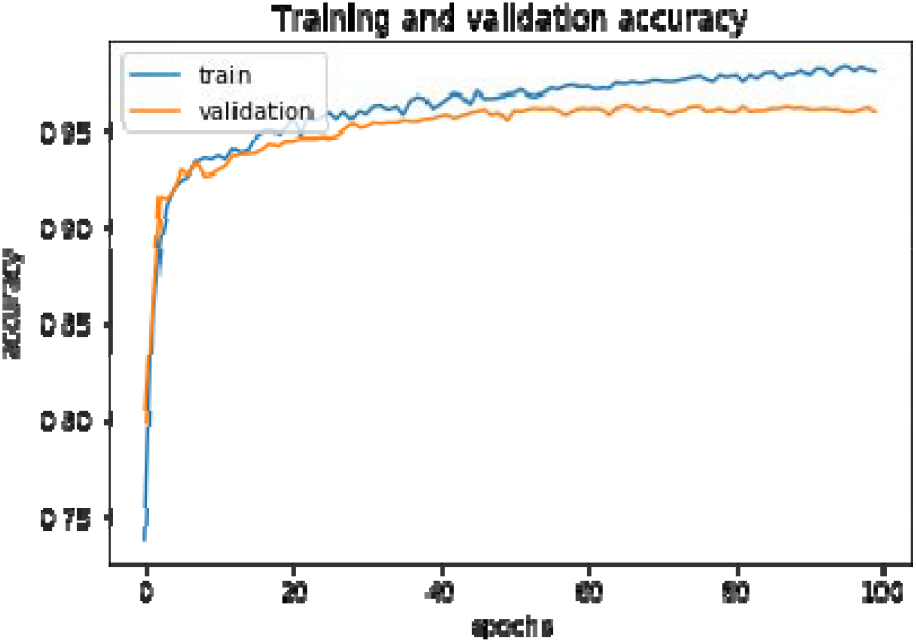
fusion model accuracy

**Figure 23.**
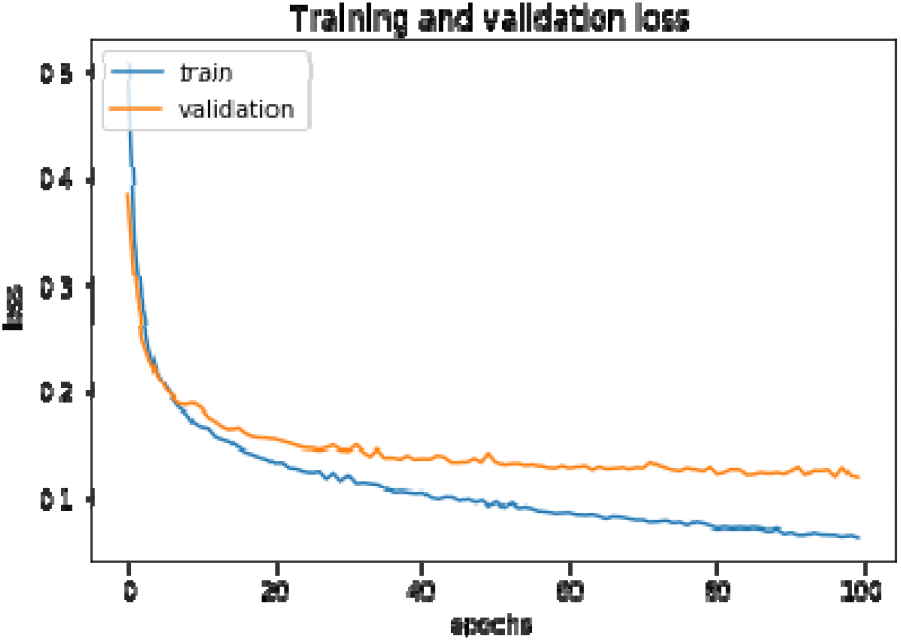
fusion model

## 5. Summary

As the large demand for new coronary pneumonia covid19 image recognition samples, the recognition accuracy is not ideal, this paper designs a new coronary pneumonia positive recognition method based on small samples, and uses image data enhancement to expand a large number of samples, solving the problem of large sample demand. Through a series of operations such as model migration, model fusion, and model fine-tuning, the recognition accuracy is improved. The experimental results show that the recognition accuracy of this experiment is about 96%, which has a high reference value for the diagnosis of new coronary pneumonia, and proves the effectiveness of the method in this paper.

## Data Availability

The data used to support the findings of this study are included within the article.

https://datadryad.org/stash/dataset/doi:10.5061/dryad.t1g1jwt1d?

## Data Availability

The data used to support the findings of this study are included within the article.

## Acknowledgments

The work of Dongsheng Ji, Yanzhong Zhao, Zhujun Zhang,and Qianchuan Zhao was supported by the Research and innovation ability improvement project of Gansu Provincial Education Department.

The project under grant 2019A-030.

